# Dietary Sodium and Potassium Patterns in Adults with Food Insecurity in the Context of Hypertension Risk

**DOI:** 10.1101/2024.02.01.24302167

**Authors:** Elizabeth A. Onugha, Ankona Banerjee, Horahenage D. Vimalajeewa, Kenneth J. Nobleza, Duc T. Nguyen, Susan B. Racette, Jayna M. Dave

**Affiliations:** Department of Pediatrics, Baylor College of Medicine, Houston, Texas; Department of Statistics, University of Nebraska, Lincoln, Nebraska; College of Health Solutions, Arizona State University, Phoenix, Arizona; USDA/ARS Children’s Nutrition Research Center

**Keywords:** Hypertension, food insecurity, dietary sodium, dietary potassium, NHANES

## Abstract

**Background:** Food insecurity (FIS), characterized by the lack of consistent access to nutritious food, is associated with hypertension and adverse health outcomes. Despite evidence of a higher prevalence of hypertension (HTN) in patients living with FIS, there is limited data exploring the underlying mechanism.

**Methods:** We conducted a cross-sectional analysis of 17,015 adults aged 18-65 years, using dietary recall data from the National Health and Nutrition Examination Survey (2011-2018). Univariate and multivariable analyses were used to examine the association between FIS, HTN, and dietary sodium and potassium levels.

**Results:** Individuals reporting FIS had a significantly lower mean intake of potassium (2.5±0.03 gm) compared to those in food-secure households (2.74±0.02 gm). No significant difference was found in the mean dietary sodium intake based on food security status. Non-Hispanic Black participants showed a high prevalence of HTN and FIS. While Non-Hispanic White and Hispanic participants had a high prevalence of FIS, it did not appear to influence their risk of HTN.

**Conclusions:** Adults with FIS and HTN were more likely to report a lower dietary potassium intake. Increasing access to healthy foods, particularly potassium-rich foods, for individuals facing FIS, may contribute to reducing the HTN prevalence and improving cardiovascular outcomes.

## Introduction

Hypertension is a significant public health issue, affecting 116 million adults in the United States.^1^ Various social determinants of health (SDoH) associated with lower socioeconomic status, including lower income level, educational attainment, employment status and opportunities, and adverse environmental factors, are associated with hypertension and cardiovascular disease (CVD) risks.^2^ Individuals in low-income areas experience higher rates of hypertension, uncontrolled hypertension and an elevated risk of fatal CVD complications,^3^ highlighting the impact of SDoH on hypertension prevalence and outcomes.

Food insecurity, one of many SDoH, is a growing public health concern.^4^ It is linked to increased hypertension risk and adverse health outcomes.^5^ US adults living below the federal poverty level exhibit a higher prevalence of food insecurity.^3^ Alarmingly, 84% of households served by the food banks in the Feeding America network experienced food insecurity, with 58% of them having a family member with hypertension.^6^ Studies reveal that individuals with food insecurity are less likely to have their hypertension under control.^3^ Addressing food insecurity alongside adopting a healthy lifestyle can significantly reduce hypertension prevalence.^7,8^ However, limited income and food insecurity pose significant barriers to implementing these positive changes.

Several pathways link food insecurity to hypertension,^9^ including stress response to poverty,^10^ suboptimal dietary quality, ^11,12^ and obesity.^13^ Individuals facing food insecurity often resort to unhealthy eating patterns characterized by inexpensive and highly processed foods that are high in sodium and nutritionally deficient.^11,12^ High sodium and low potassium intake, major risk factors of hypertension, are associated with diets low in vegetables, fruits, nuts, and omega-3 fatty acids, and high in processed meats. ^14^ Food-insecure households often face challenges in paying bills, making trade-offs between food and other financial obligations. ^6, 15^

Despite the clear relationship between poor diets and hypertension, limited studies explore the impact of food insecurity on hypertension. However, studies indicating an association between poor diet quality, chronic disease risk, and cardiovascular health in individuals with food insecurity suggest a potential connection.^5,7,10^ Individuals with lower income and food insecurity tend to consume more highly palatable salty meals,^11, 12^ reinforcing a plausible link between food insecurity and hypertension is plausible.

Our study aims to assess the associations between food insecurity, hypertension, and dietary sodium and potassium in a nationally representative cohort of adults. We hypothesize that adults experiencing food insecurity and hypertension would exhibit higher dietary sodium and lower potassium intakes compared to adults who are food secure and have normal blood pressure (BP).

## Methods

### Study Population

We conducted a cross sectional secondary analysis using data from the National Health and Nutrition Examination Survey (NHANES) from 2011-2018. NHANES is a nationally representative survey assessing the nutrition and health condition of noninstitutionalized individuals through interviews and physical examinations, conducted by the National Center for Health Statistics (NCHS).^16^ Ethical approval was exempt for this study because we used publicly available, de-identified data.

Our analysis included adults between the ages of 18 and 65 years. We excluded pregnant women and participants with incomplete BP and food security screening data.

### Blood pressure status

Blood pressure was classified into normal BP, elevated BP and hypertension based on the 2017 American College of Cardiology (ACC)/American Heart Association (AHA) Task Force guidelines.^17^ Participants who reported use of antihypertensive medications were categorized as having hypertension.

### Food security status

Food security was determined using the 18-item USDA’s Food Security Module and categorized into one of four food security categories (i.e., high, marginal, low, and very low) based on NHANES documentation.^18^

### Dietary recall data

Dietary intake of sodium and potassium was evaluated via two 24-hour dietary recall interviews. Detailed information on 24-hour dietary recall procedures can be found elsewhere.^19^

### Demographics and SDoH factors

Demographic information included age at the time of the interview, sex, and self-reported race/ethnicity. The income-to-poverty ratio variable was utilized to assess participants’ socioeconomic status adjusted for family size, as established by the U.S. Census Bureau, and divided into 4 distinct categories: Family income ≤ 1.3 (poorest); between 1.3 and 1.85; >1.85 of the poverty threshold and N/A, missing or incomplete data.

### Physical Activity

We estimated the weekly number of minutes spent performing moderate and vigorous physical activities. Using the Physical Activity questionnaire, participants were categorized into three groups (high or recommended, intermediate, and low physical activity) based the CDC’s recommended level of physical activity.^20^

### Statistical Analyses

Descriptive data were reported as proportion and 95% confidence interval for categorical variables, and as mean (± standard error, SE) for continuous variables. Patient characteristics were compared based on BP status (normal, elevated, or hypertension) and food security status using the chi-squared test for the categorical variables and the t-test for the continuous variables, considering sampling weights. To address missing data and imbalances in the categories, high food security and marginal food security were merged as one category of food security; low and very low food security were merged to classify food insecurity. Age was categorized into five-year intervals, as suggested by Diaz et al.^21^ Participants aged 18 to 29 years were grouped together, considering the lower prevalence of primary hypertension in individuals under 30.

Univariate logistic regression models were used to investigate variables associated with hypertension and food insecurity. Quasi-binomial distribution was applied in univariate and multivariable logistic regression to explore the relationship between food security, hypertension, dietary sodium and potassium intake, socioeconomic status, and health disparities.

Outcome measures included food security status and hypertension, with each serving as predictor and outcome variable, depending on the analysis. Predictor variables included race/ethnicity, sex, food security status, body mass index (BMI), diabetes, kidney disease, sodium and potassium intake, age, physical activity, and family income. Separate logistic regression models for survey data were fitted to identify risk factors associated with hypertension or food insecurity. All analyses were conducted using R version 4.3.1 (R Core team, Vienna, Austria). All tests were two-sided and a p-value <0.05 was considered statistically significant.

We presented results as unweighted counts (actual numbers of participants sampled) and weighted estimates. The NCHS sample weights in NHANES account for the complex survey design (including oversampling), survey non-response, and post-stratification adjustment to match total population counts from the Census Bureau, ensuring representative estimates of the U.S. civilian noninstitutionalized population.^18^

## Results

### Participant Characteristics

During the study period, 39,156 individuals completed the NHANES surveys. The final dataset for our analysis included 17,015 adults aged 18 to 65 years (Figure 1). Of these 15,633 participants completed the 24-hour dietary recall.

**Figure 1:**
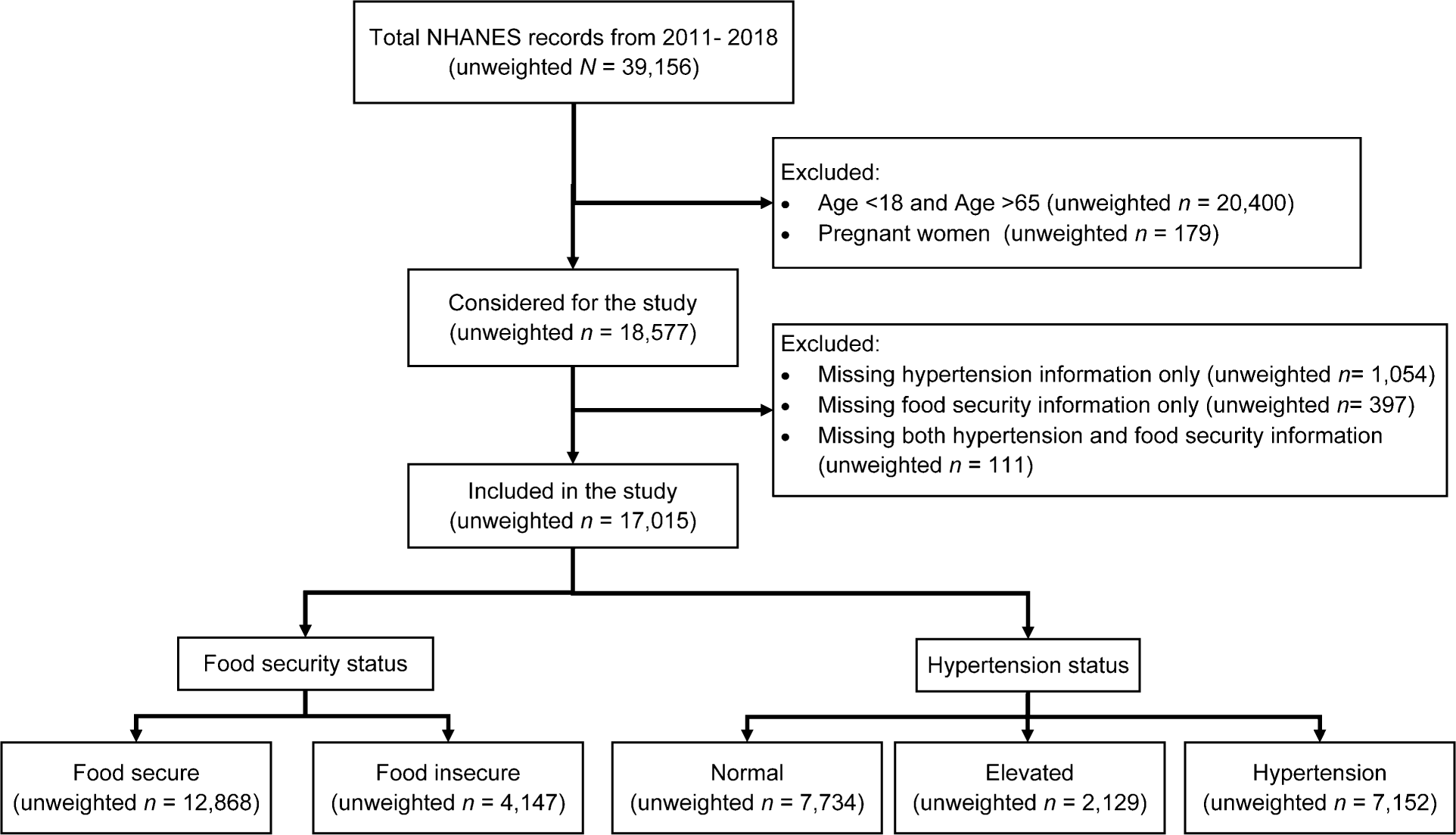
Participant Selection.

### Hypertension as an outcome

Weighted estimates revealed distinctive characteristics between participants with hypertension and those with normal BP. The hypertension group was significantly older, more likely male, and obese and had a significantly higher dietary sodium intake compared to the normal BP group (Table 1). The prevalence of hypertension in US adults across different time periods, and in age, sex, and race-ethnicity subgroups is presented in Table S1.

**Table 1:**
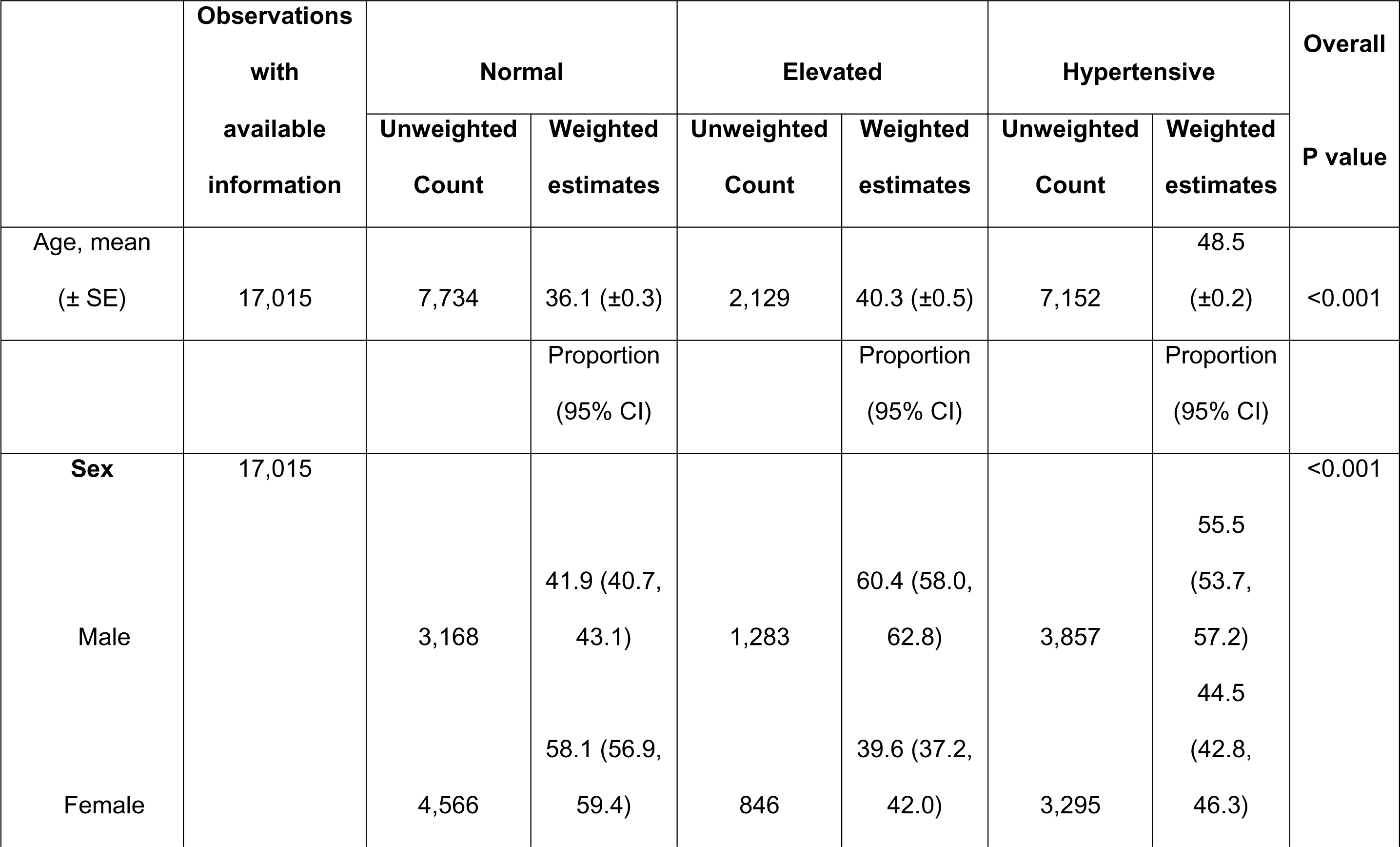
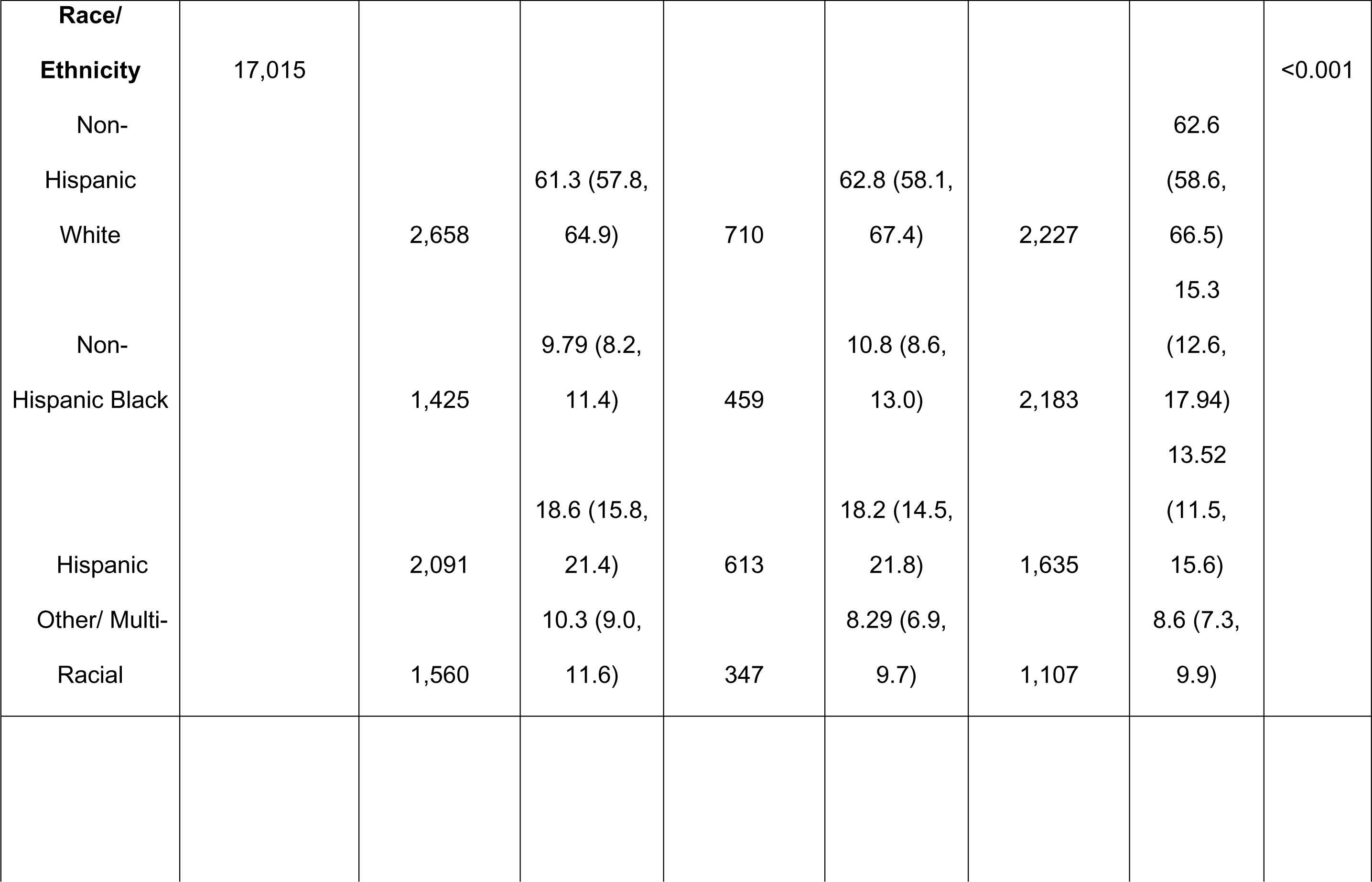
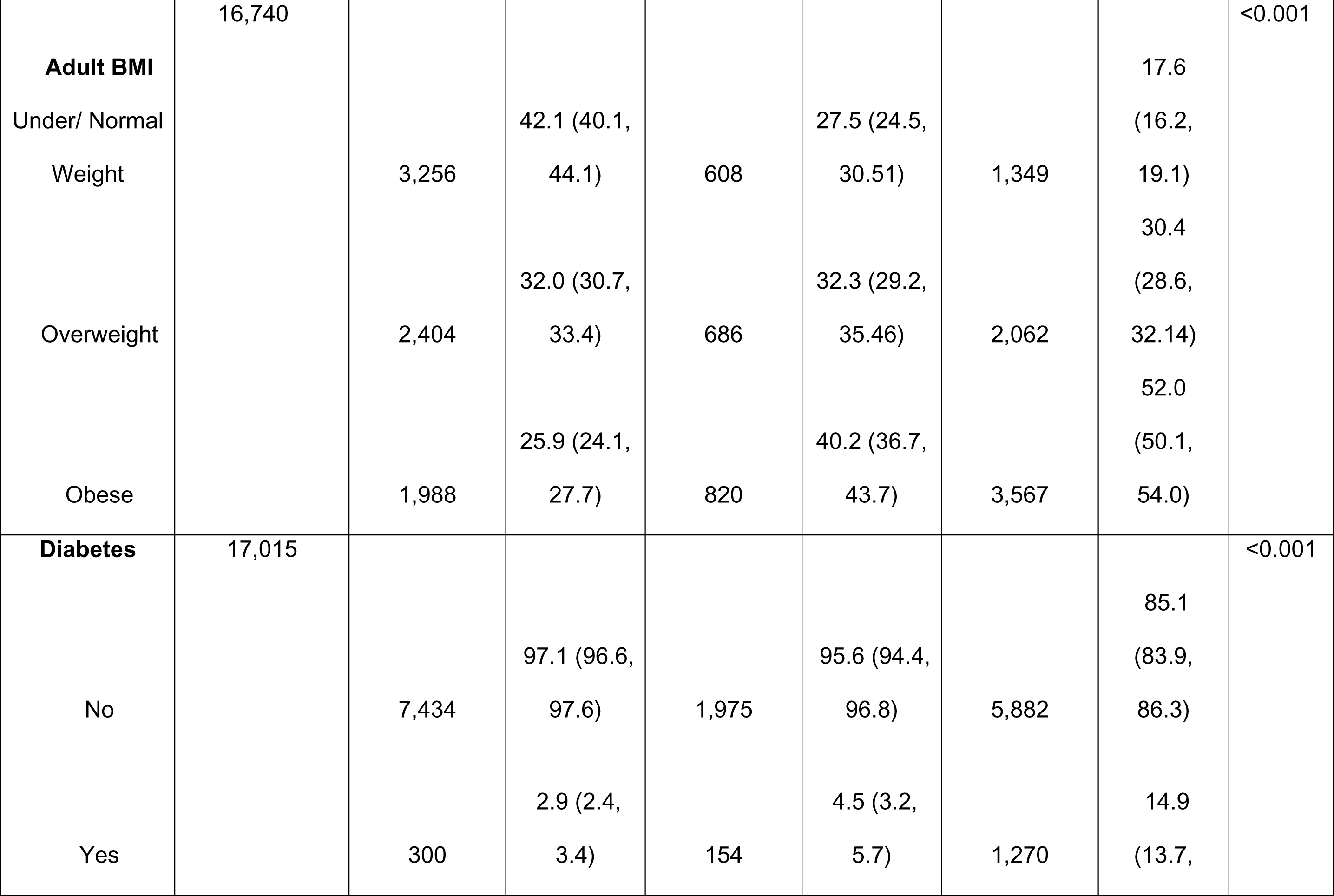
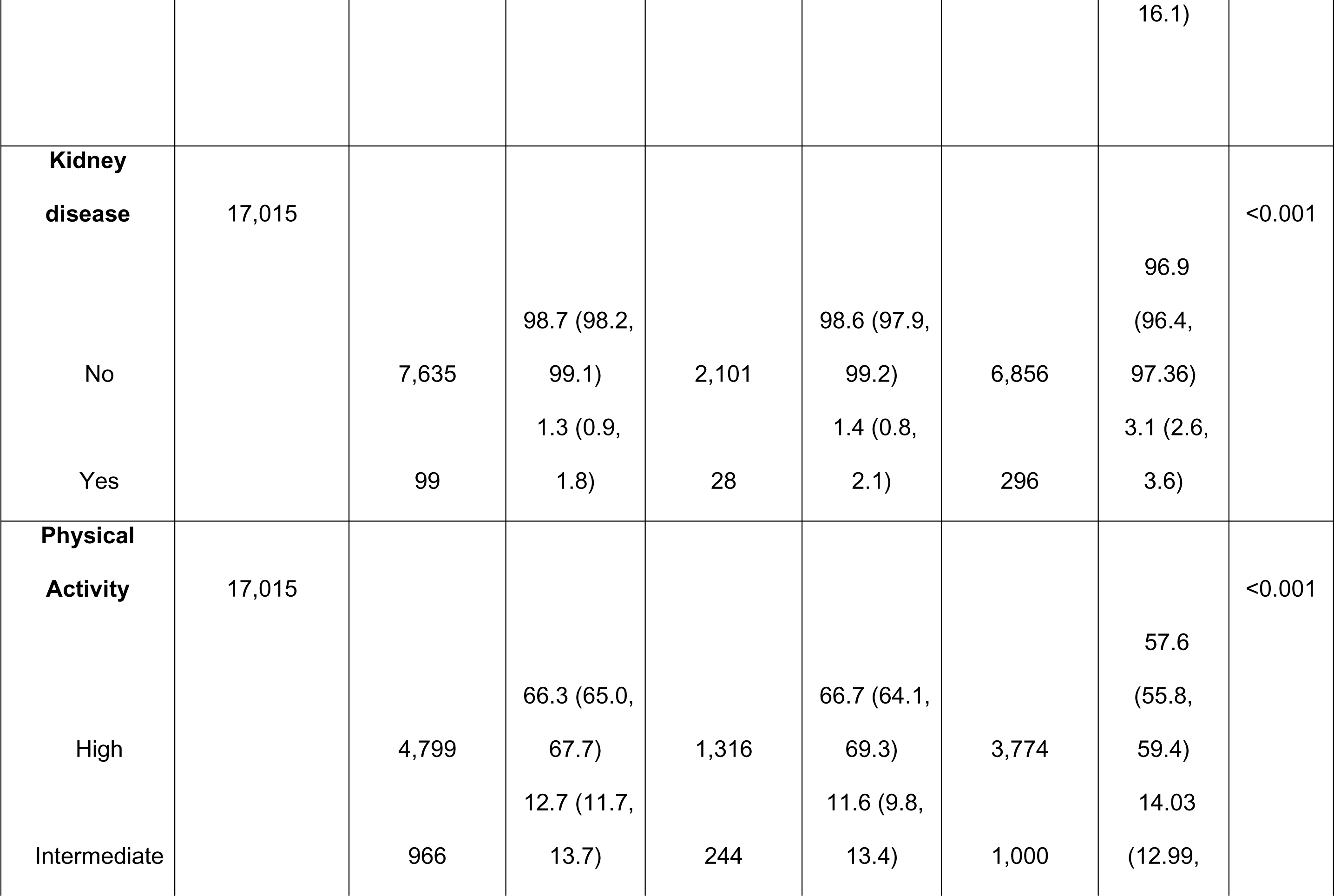
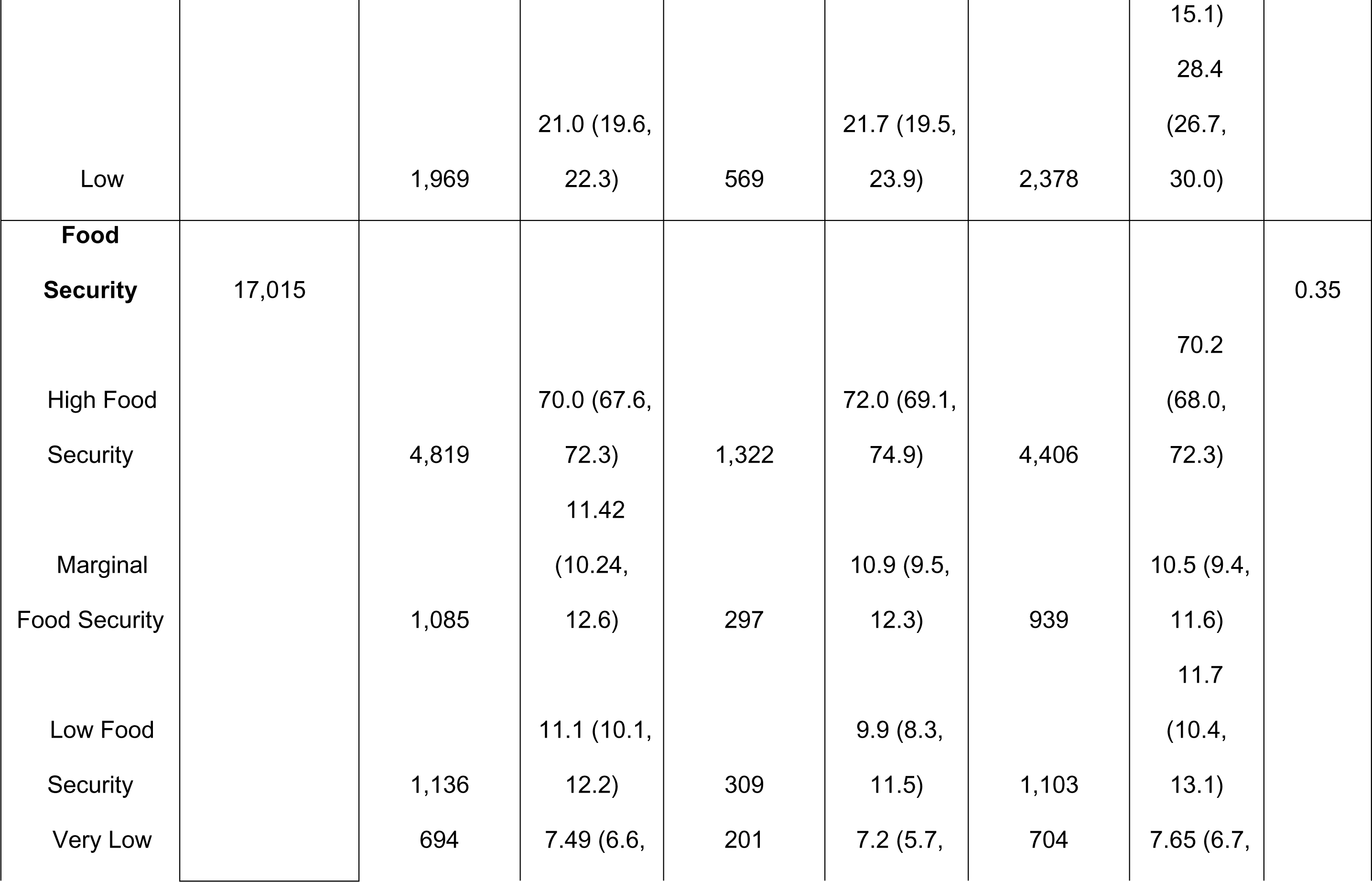
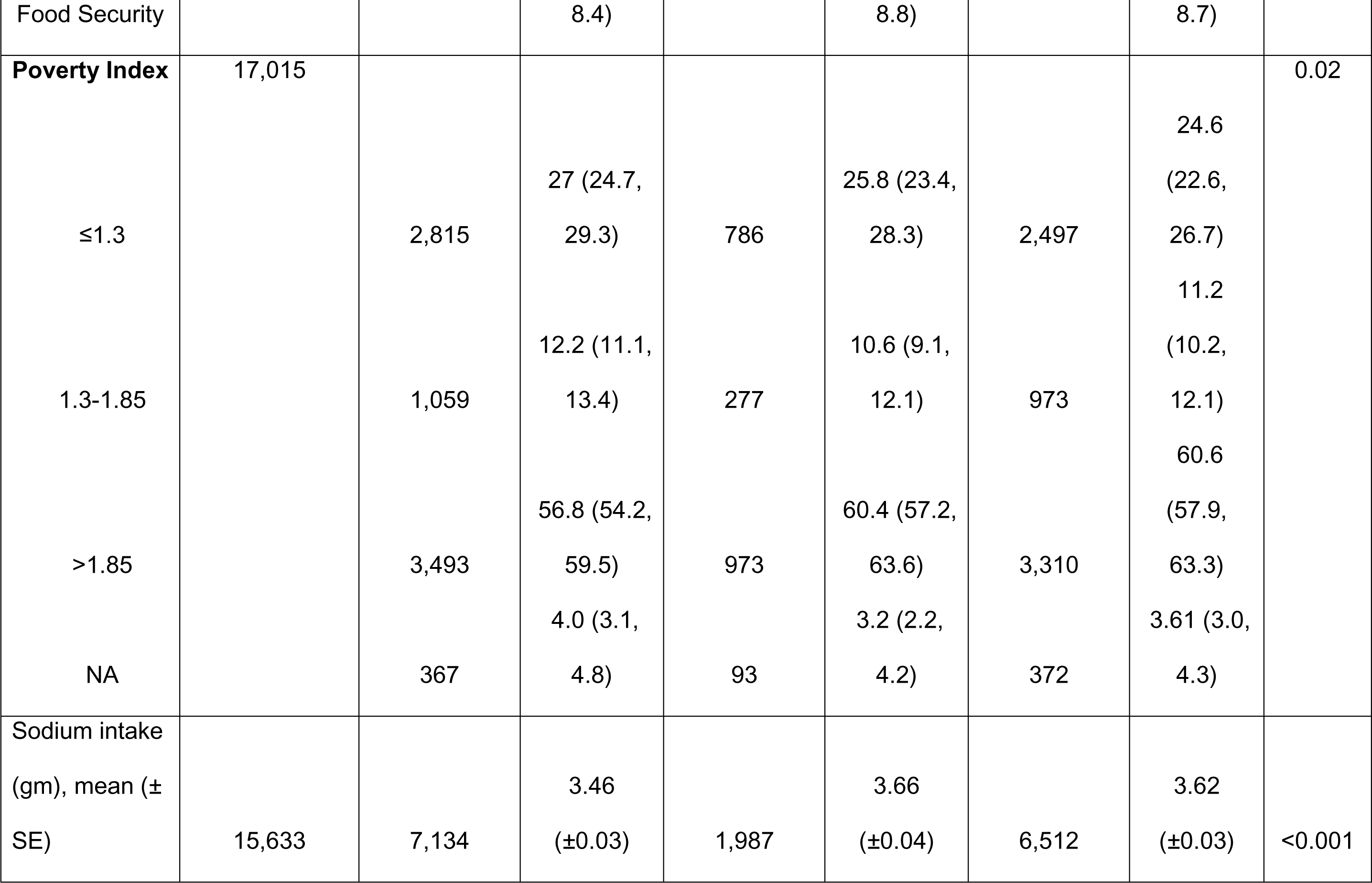
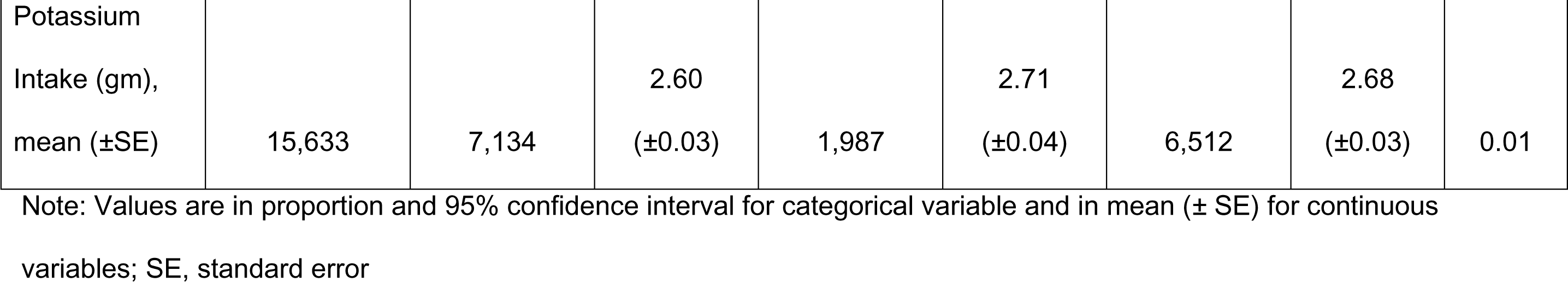
Characteristics by blood pressure status for NHANES 2011-2018 participants.

We observed a dose-response relationship between hypertension and age categories. Using the 18-29.9 group as a reference, we found a stepwise increase in hypertension risk with each older age group. For example, the odds of hypertension were 1.7 (1.3-2.2) for the 30-34.9 age group, 3.8 (2.9-4.9) for the 40-44.9 age group, reaching 15.6 (12.2-19.8) in the 60-64.9 age group (Table 2).

**Table 2:**
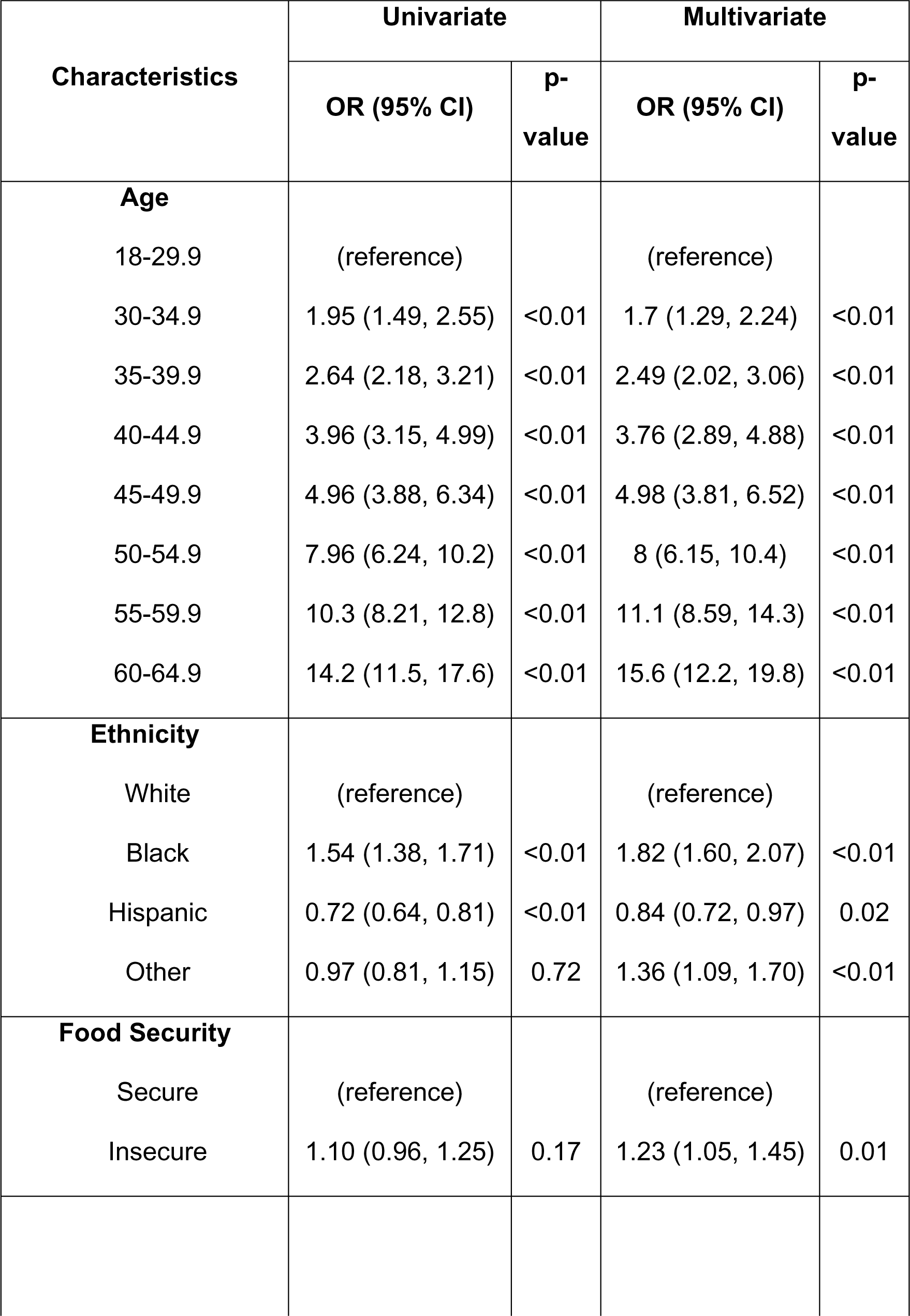
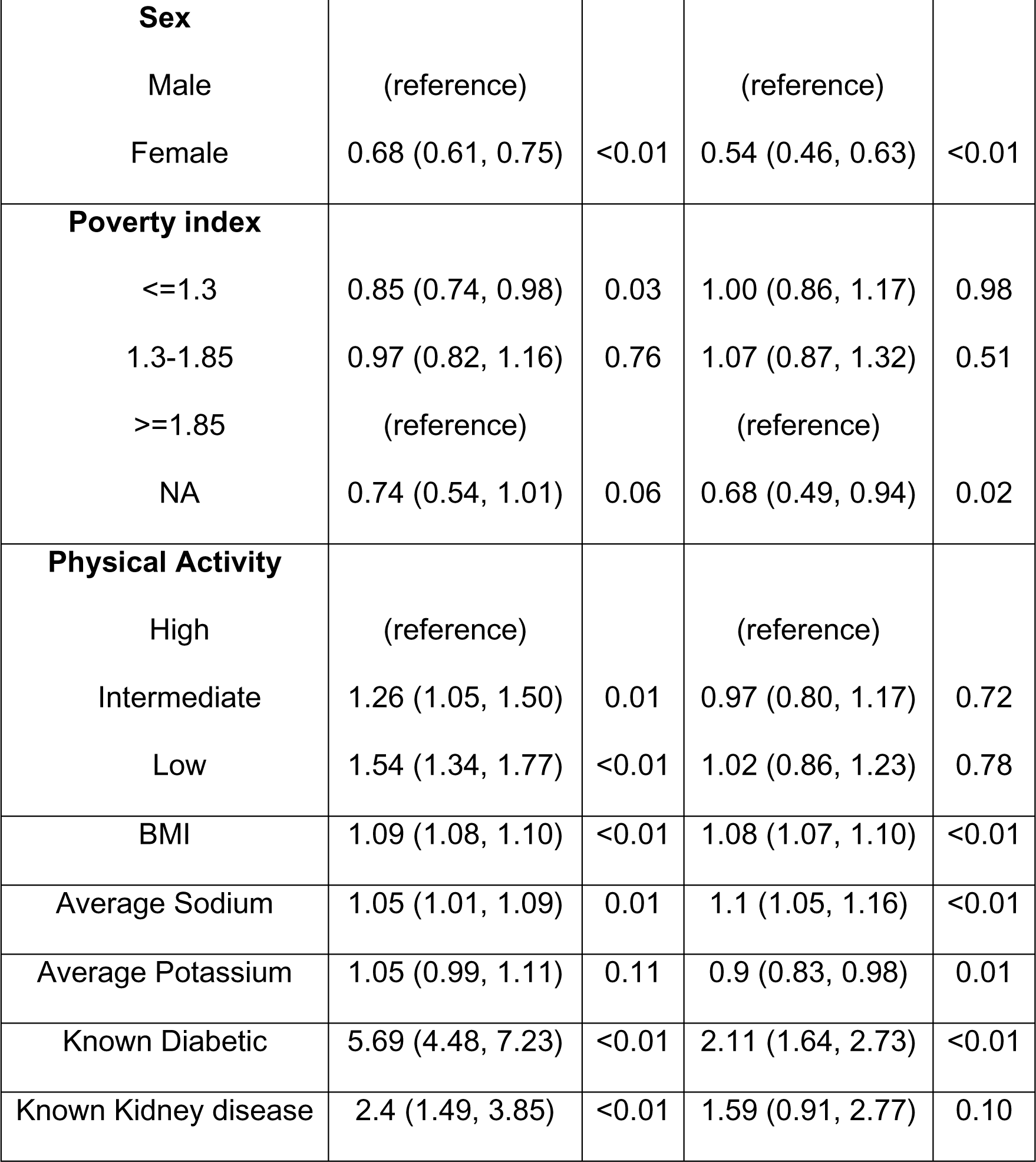
Logistic regression with hypertension as an outcome.

| Characteristics | Univariate |  | Multivariate |  |
| --- | --- | --- | --- | --- |
|  | OR (95% CI) | p-value | OR (95% CI) | p-value |
| **Age** |  |  |  |  |
| 18-29.9 | (reference) |  | (reference) |  |
| 30-34.9 | 1.95 (1.49, 2.55) | <0.01 | 1.7 (1.29, 2.24) | <0.01 |
| 35-39.9 | 2.64 (2.18, 3.21) | <0.01 | 2.49 (2.02, 3.06) | <0.01 |
| 40-44.9 | 3.96 (3.15, 4.99) | <0.01 | 3.76 (2.89, 4.88) | <0.01 |
| 45-49.9 | 4.96 (3.88, 6.34) | <0.01 | 4.98 (3.81, 6.52) | <0.01 |
| 50-54.9 | 7.96 (6.24, 10.2) | <0.01 | 8 (6.15, 10.4) | <0.01 |
| 55-59.9 | 10.3 (8.21, 12.8) | <0.01 | 11.1 (8.59, 14.3) | <0.01 |
| 60-64.9 | 14.2 (11.5, 17.6) | <0.01 | 15.6 (12.2, 19.8) | <0.01 |
| **Ethnicity** |  |  |  |  |
| White | (reference) |  | (reference) |  |
| Black | 1.54 (1.38, 1.71) | <0.01 | 1.82 (1.60, 2.07) | <0.01 |
| Hispanic | 0.72 (0.64, 0.81) | <0.01 | 0.84 (0.72, 0.97) | 0.02 |
| Other | 0.97 (0.81, 1.15) | 0.72 | 1.36 (1.09, 1.70) | <0.01 |
| **Food Security** |  |  |  |  |
| Secure | (reference) |  | (reference) |  |
| Insecure | 1.10 (0.96, 1.25) | 0.17 | 1.23 (1.05, 1.45) | 0.01 |
| **Sex** |  |  |  |  |
| Male | (reference) |  | (reference) |  |
| Female | 0.68 (0.61, 0.75) | <0.01 | 0.54 (0.46, 0.63) | <0.01 |
| **Poverty index** |  |  |  |  |
| <=1.3 | 0.85 (0.74, 0.98) | 0.03 | 1.00 (0.86, 1.17) | 0.98 |
| 1.3-1.85 | 0.97 (0.82, 1.16) | 0.76 | 1.07 (0.87, 1.32) | 0.51 |
| >=1.85 | (reference) |  | (reference) |  |
| NA | 0.74 (0.54, 1.01) | 0.06 | 0.68 (0.49, 0.94) | 0.02 |
| **Physical Activity** |  |  |  |  |
| High | (reference) |  | (reference) |  |
| Intermediate | 1.26 (1.05, 1.50) | 0.01 | 0.97 (0.80, 1.17) | 0.72 |
| Low | 1.54 (1.34, 1.77) | <0.01 | 1.02 (0.86, 1.23) | 0.78 |
| BMI | 1.09 (1.08, 1.10) | <0.01 | 1.08 (1.07, 1.10) | <0.01 |
| Average Sodium | 1.05 (1.01, 1.09) | 0.01 | 1.1 (1.05, 1.16) | <0.01 |
| Average Potassium | 1.05 (0.99, 1.11) | 0.11 | 0.9 (0.83, 0.98) | 0.01 |
| Known Diabetic | 5.69 (4.48, 7.23) | <0.01 | 2.11 (1.64, 2.73) | <0.01 |
| Known Kidney disease | 2.4 (1.49, 3.85) | <0.01 | 1.59 (0.91, 2.77) | 0.10 |

Several factors including sex, age, race/ethnicity, and food security status were significantly associated with an increased likelihood of hypertension (Table 2). Non-Hispanic Blacks had higher odds of hypertension compared to Non-Hispanic Whites (OR=1.82, 95% CI: 1.60-2.07, p<0.001). Participants reporting food insecurity also had higher odds of hypertension (OR=1.23, 95% CI: 1.05-1.45, p=0.01).

In terms of dietary habits, the hypertension group consumed slightly higher levels of sodium (3.62 gm, SE=0.03) (p=0.01) compared to the normal BP group, while there was no significant difference in potassium intake between the two groups (Table 2).

The study highlighted the positive associations between increasing BMI and higher dietary sodium intake with hypertension (Table 3). For each unit increase in BMI, the odds of hypertension increased significantly by 8% (OR = 1.08; 95% CI: 1.07-1.10, p <0.01). Similarly, with every one-gram increase in dietary sodium intake, the odds of hypertension increased by 1% (p< 0.01). Conversely, with every one-gram increase in dietary potassium intake, the odds of hypertension decreased by 9% (p<0.01). The model’s AUC values of 0.81 suggest a relatively strong discriminative ability between the hypertensive and non-hypertensive groups.

**Table 3:** Characteristics by food security status for NHANES 2011-2018 participants.

|  | Unweighted | Food Secure |  | Food Insecure |  | P<br>value |
| --- | --- | --- | --- | --- | --- | --- |
|  | count with<br>available<br>data | Unweighted<br>count | Weighted<br>estimates | Unweighted<br>count | Weighted<br>estimates |  |
| Age, mean (±SE) | 17,015 | 12,868 | 42.1 (±0.3) | 4,147 | 39.3 (±0.4) | <0.001 |
|  |  |  | Proportion<br>(95% CI) |  | Proportion<br>(95% CI) |  |
| **Sex** | 17,015 |  |  |  |  | 0.02 |
| Male |  | 6,333 | 50.2 (49.3, 51.1) | 1,975 | 47.7 (45.8, 49.5) |  |
| Female |  | 6,535 | 49.8 (48.9, 50.6) | 2,172 | 52.3 (50.5, 54.1) |  |
| **Race/ Ethnicity** | 17,015 |  |  |  |  | <0.001 |
| Non- Hispanic White |  | 4,386 | 65.4 (61.9, 68.9) | 1,209 | 47.2 (42.4, 52.0) |  |
| Non-Hispanic Black |  | 2,935 | 10.8 (8.9, 12.7) | 1,132 | 17.7 (14.7, 20.7) |  |
| Hispanic |  | 2,939 | 14.0 (11.7, 16.3) | 1,400 | 27.5 (23.5, 31.4) |  |
| Other/ Multi-Racial |  | 2,608 | 9.8 (8.5, 11.0) | 2,608 | 7.7 (6.2, 9.2) |  |
| **Adult BMI** | 16,740 |  |  |  |  | <0.001 |
| Under/ Normal Weight |  | 4,114 | 31.2 (29.5, 33.0) | 1,099 | 27.1 (24.6, 29.7) |  |
| Overweight |  | 4,000 | 32.3 (30.9, 33.6) | 1,152 | 27.6 (25.8, 29.5) |  |
| Obese |  | 4,557 | 36.5 (34.7, 38.4) | 1,818 | 45.3 (42.8, 47.7) |  |
| **Hypertension** | 17,015 |  |  |  |  | 0.18 |
| No |  | 7,523 | 60.5 (59.2, 61.9) | 2,340 | 58.9 (56.5, 61.2) |  |
| Yes |  | 5,345 | 39.5 (38.2, 40.8) | 1,807 | 41.2 (38.8, 43.5) |  |
| **Diabetes** | 17,015 |  |  |  |  | 0.001 |
| No |  | 11,668 | 92.6 (92.0, 93.1) | 3,623 | 90.3 (89.1, 91.6) |  |
| Yes |  | 1,200 | 7.5 (6.9, 8.0) | 524 | 9.7 (8.3, 10.9) |  |
| **Kidney disease** | 17,015 |  |  |  |  | <0.001 |
| No |  | 12,603 | 98.2 (97.9, 98.6) | 3,989 | 96.8 (96.1, 97.4) |  |
| Yes |  | 265 | 1.8 (1.4, 2.2) | 158 | 3.25 (2.6, 3.9) |  |
| **Physical Activity** |  |  |  |  |  |  |
| High | 17,015 | 7,584 | 63.6 (62.5, 64.) | 2,305 | 60.0 (58.0, 62.0) | <0.001 |
| Intermediate |  | 1,755 | 13.7 (12.9, 14.5) | 455 | 10.5 (9.3, 11.7) |  |
| Low |  | 3,529 | 22.74 (21.7, 23.8) | 1,387 | 29.5 (27.8, 31.3) |  |
| **Poverty** | 17,015 |  |  |  |  | <0.001 |
| Poverty <=1.3 |  | 3,469 | 18.5 (16.8, 20.2) | 2,629 | 58.0 (52.4, 63.6) |  |
| 1.3-1.85 |  | 1,683 | 10.7 (9.8, 11.6) | 626 | 15.4 (13.2, 17.6) |  |
| >1.85 |  | 7,053 | 67.0 (64.9, 69.2) | 723 | 23.2 (20.8, 25.5) |  |
| NA |  | 663 | 3.4 (2.6, 4.2) | 169 | 3.4 (2.6, 4.2) |  |
| Sodium Intake (gm), mean<br>(±SE) | 15,633 | 11,796 | 3.6 (±0.02) | 3,837 | 3.42 (±0.04) | <0.001 |
| Potassium Intake (gm), mean<br>(±SE) | 15,633 | 11,796 | 2.7 (±0.02) | 3,837 | 2.41 (±0.03) | <0.001 |
| Note: Values are in proportion and 95% confidence interval for categorical variable and in mean (± SE) for continuous variables; SE, standard error |  |  |  |  |  |  |

### Food Insecurity as an outcome

Significant differences in demographic, lifestyle, and health factors were observed between the food secure and food insecure groups. Participants reporting food insecurity were younger than food secure participants were (average age of 39.29 ± 0.35 years vs. 42.10 ± 0.27 years) (p < 0.01) based on weighted estimates. Additionally, food insecure participants were predominantly female (52%) and obese (45%) compared to food-secure counterparts (Table 3).

Ethnic/racial minorities were overrepresented in the group experiencing food insecurity. In multivariate analysis, Non-Hispanic Black (OR=1.33; 95% CI: 1.05-1.68, p=0.02) and Hispanic: OR=1.78; 95% CI: 1.37-2.17, p<0.01) participants were significantly more likely to experience food insecurity compared to their White counterparts (Table 4).

**Table 4.**
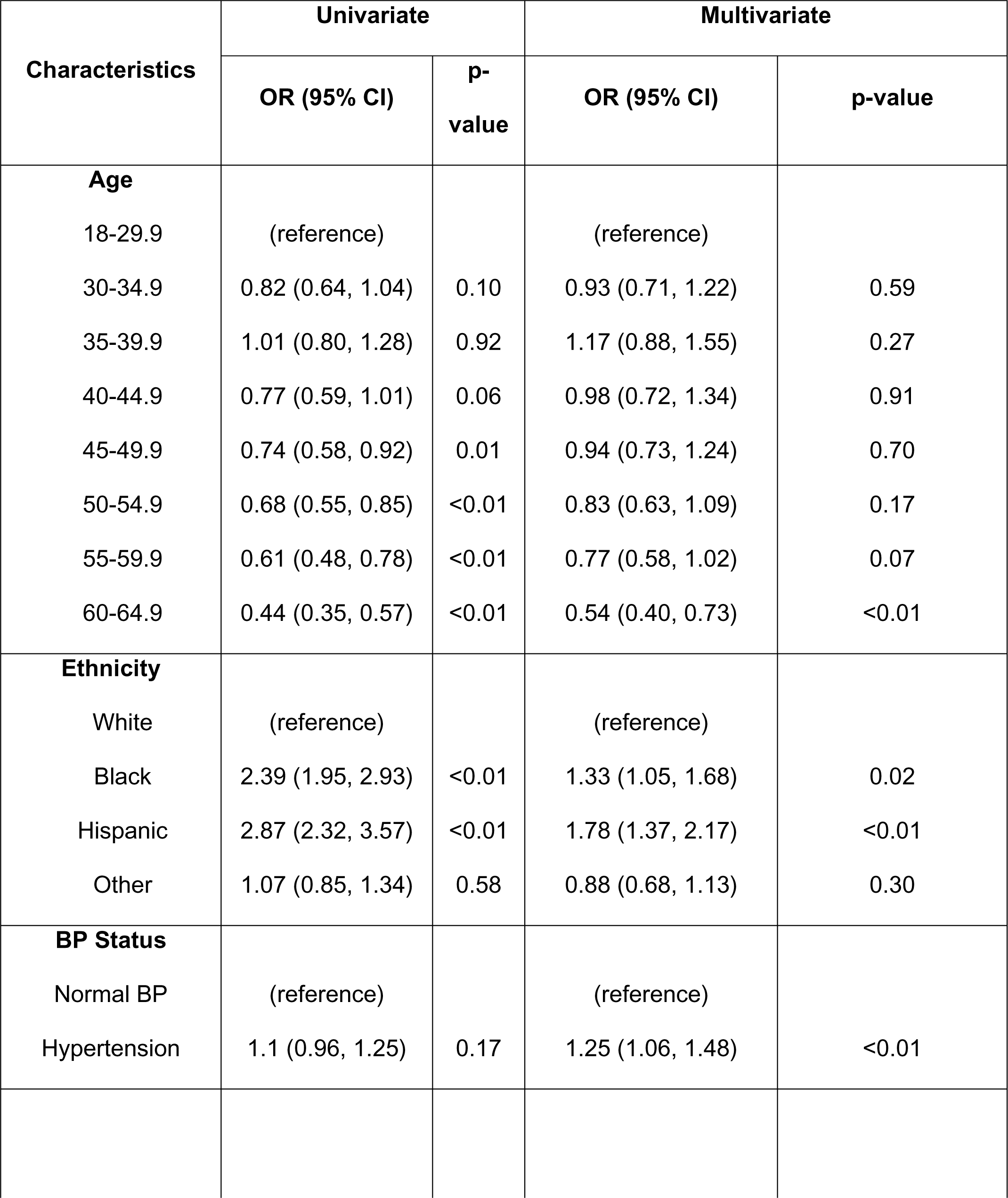
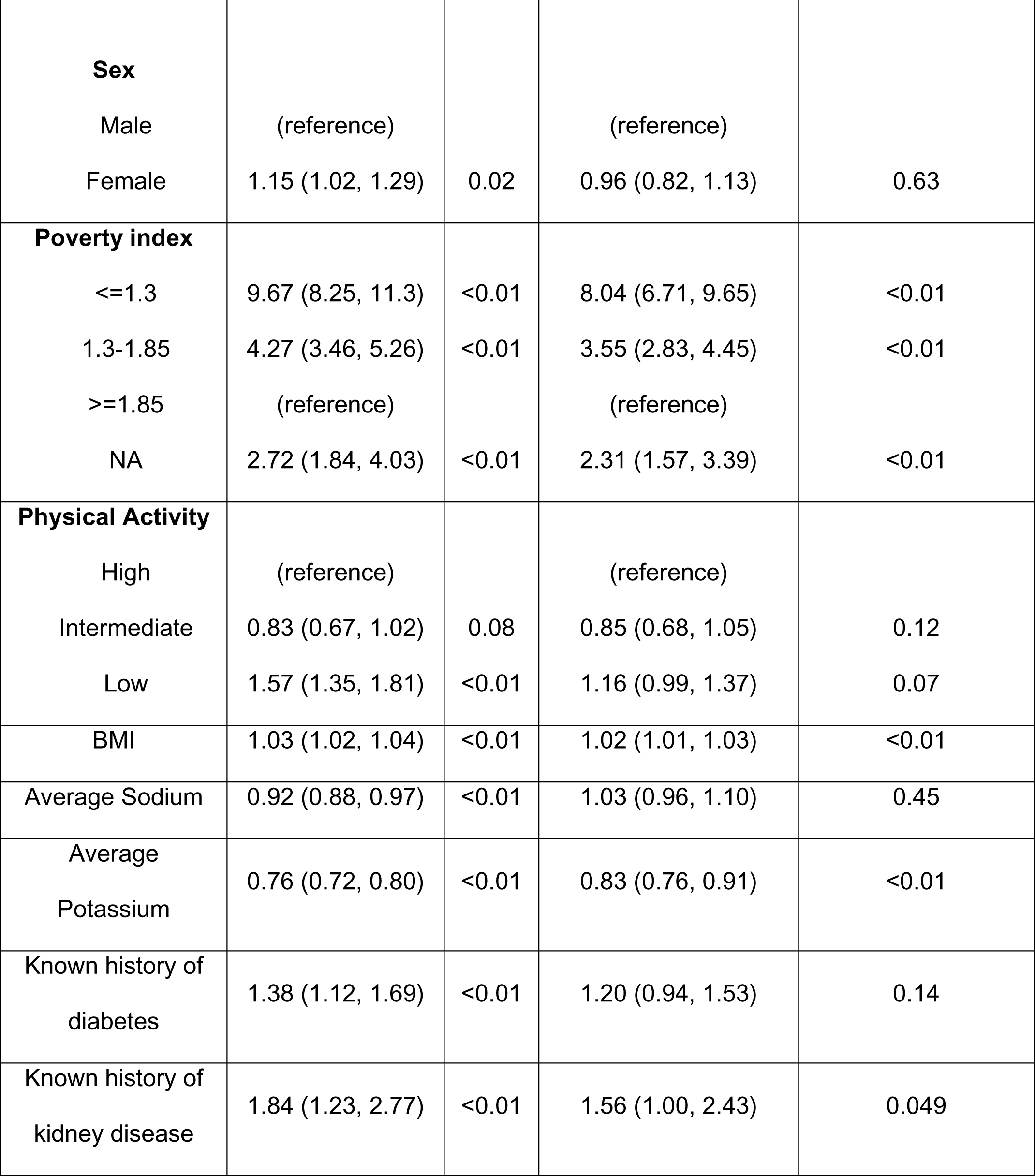
Logistic regression with food insecurity as an outcome.

| **Characteristics** | **Univariate** |  | **Multivariate** |  |
| --- | --- | --- | --- | --- |
|  | **OR (95% CI)** | **p-value** | **OR (95% CI)** | **p-value** |
| **Age** |  |  |  |  |
| 18-29.9 | (reference) |  | (reference) |  |
| 30-34.9 | 0.82 (0.64, 1.04) | 0.10 | 0.93 (0.71, 1.22) | 0.59 |
| 35-39.9 | 1.01 (0.80, 1.28) | 0.92 | 1.17 (0.88, 1.55) | 0.27 |
| 40-44.9 | 0.77 (0.59, 1.01) | 0.06 | 0.98 (0.72, 1.34) | 0.91 |
| 45-49.9 | 0.74 (0.58, 0.92) | 0.01 | 0.94 (0.73, 1.24) | 0.70 |
| 50-54.9 | 0.68 (0.55, 0.85) | <0.01 | 0.83 (0.63, 1.09) | 0.17 |
| 55-59.9 | 0.61 (0.48, 0.78) | <0.01 | 0.77 (0.58, 1.02) | 0.07 |
| 60-64.9 | 0.44 (0.35, 0.57) | <0.01 | 0.54 (0.40, 0.73) | <0.01 |
| **Ethnicity** |  |  |  |  |
| White | (reference) |  | (reference) |  |
| Black | 2.39 (1.95, 2.93) | <0.01 | 1.33 (1.05, 1.68) | 0.02 |
| Hispanic | 2.87 (2.32, 3.57) | <0.01 | 1.78 (1.37, 2.17) | <0.01 |
| Other | 1.07 (0.85, 1.34) | 0.58 | 0.88 (0.68, 1.13) | 0.30 |
| **BP Status** |  |  |  |  |
| Normal BP | (reference) |  | (reference) |  |
| Hypertension | 1.1 (0.96, 1.25) | 0.17 | 1.25 (1.06, 1.48) | <0.01 |
| **Sex** |  |  |  |  |
| Male | (reference) |  | (reference) |  |
| Female | 1.15 (1.02, 1.29) | 0.02 | 0.96 (0.82, 1.13) | 0.63 |
| **Poverty index** |  |  |  |  |
| <=1.3 | 9.67 (8.25, 11.3) | <0.01 | 8.04 (6.71, 9.65) | <0.01 |
| 1.3-1.85 | 4.27 (3.46, 5.26) | <0.01 | 3.55 (2.83, 4.45) | <0.01 |
| >=1.85 | (reference) |  | (reference) |  |
| NA | 2.72 (1.84, 4.03) | <0.01 | 2.31 (1.57, 3.39) | <0.01 |
| **Physical Activity** |  |  |  |  |
| High | (reference) |  | (reference) |  |
| Intermediate | 0.83 (0.67, 1.02) | 0.08 | 0.85 (0.68, 1.05) | 0.12 |
| Low | 1.57 (1.35, 1.81) | <0.01 | 1.16 (0.99, 1.37) | 0.07 |
| BMI | 1.03 (1.02, 1.04) | <0.01 | 1.02 (1.01, 1.03) | <0.01 |
| Average Sodium | 0.92 (0.88, 0.97) | <0.01 | 1.03 (0.96, 1.10) | 0.45 |
| Average Potassium | 0.76 (0.72, 0.80) | <0.01 | 0.83 (0.76, 0.91) | <0.01 |
| Known history of diabetes | 1.38 (1.12, 1.69) | <0.01 | 1.20 (0.94, 1.53) | 0.14 |
| Known history of kidney disease | 1.84 (1.23, 2.77) | <0.01 | 1.56 (1.00, 2.43) | 0.049 |

Participants reporting food insecurity exhibited a significantly lower intake of dietary potassium (2.41 ± 0.03 gm) compared to food secure individuals (2.79 ± 0.02 gm); p < 0.01). Conversely, participants with food insecurity had significantly lower mean intakes of sodium (3.42 ± 0.04 gm) compared to food secure individuals (3.58 ± 0.02 gm; p < 0.01) (Table 2). Higher average potassium intake was associated with lower odds of food insecurity (OR=0.83; 95% CI: 0.76-0.91, p<0.01), while no significant association was found with average sodium intake in the multivariable analysis (Table 4).

With respect to health characteristics, participants reporting food insecurity had a higher prevalence of obesity (45.26%) compared to the food secure group (36.52%; p < 0.01). For each unit increase in BMI, the odds of food insecurity marginally increased in both univariate (OR=1.03; 95% CI: 1.02-1.04, p<0.01) and multivariable (OR=1.02; 95% CI: 1.01-1.03, p<0.01) analyses (Table 4).

While no statistically significant difference in the prevalence of hypertension between the food secure and food insecure groups, multivariable analysis revealed that participants with hypertension were more likely to experience food insecurity (OR=1.25; 95% CI: 1.06-1.48, p<0.01). The association between a reported history of diabetes and food insecurity (OR=1.38; 95% CI: 1.12-1.69, p<0.01) was not significant in the multivariable analysis. Similarly, participants with a history of kidney disease had a higher likelihood of food insecurity in the univariate analysis (OR=1.84, 95 CI: 1.23-2.77, p<0.01) and approached significance in the multivariable analysis (Table 4).

Furthermore, participants with low socioeconomic status, defined by a poverty index ≤1.3 (OR=8.04; 95% CI: 6.71-9.65, p<0.01) and 1.3-1.85 (OR=3.55; 95% CI: 2.83-4.45, p<0.01), had significantly higher odds of experiencing food insecurity than those with a poverty index ≥1.85 in multivariable analyses.

Finally, we observed that the level of physical activity appeared to influence food insecurity. In the multivariate analysis, participants who reported low activity levels demonstrated a higher likelihood of experiencing food insecurity (OR=1.16; 95% CI: 0.99-1.37, p=0.07) compared to participants with high levels of physical activity. The model’s AUC values of 0.75 suggest a relatively moderate discriminative ability between the food-secure and insecure groups.

## Discussion

This cross-sectional study explored patterns of dietary sodium and potassium intake based on food security status in relation to hypertension risk. Food insecure participants reported a lower dietary potassium intake compared to those without food insecurity, likely related to lower intake of potassium-rich foods such as fruits, vegetables, and grains, which are main components of the DASH (Dietary Approaches to Stop Hypertension) eating plan.^8^ This aligns with previous models demonstrating reduced dietary intake of vegetables and fruits as leading risk factors for cardiometabolic mortality.^22^

While individuals with hypertension had higher mean dietary sodium intake compared to individuals without hypertension, as has been previously established,^14^ no significant difference was found in dietary sodium intake based on food security status. Specifically, participants who reported food insecurity had a lower dietary sodium intake compared to those who were food secure. Dietary recall remains a valuable tool to assess sodium intake, but acknowledging potential recall bias and the inherent variability in nutrient contents of foods in the food composition tables is important. ^23^ For clinical and health-related research, exploring complementary methods (e.g. 24-hour urine collection) may be necessary to enhance accuracy and address these limitations. Given the strong link between reduced dietary potassium intake and hypertension, public health initiatives should focus on increasing access to potassium-rich foods, especially among populations at risk of food insecurity.^8^

Similar to previous reports, our study observed a significant association between food insecurity and hypertension among U.S. adults,^24,25^ reinforcing the link between food insecurity and chronic diseases like hypertension.^10^ Understanding the mechanisms contributing to hypertension risk in food insecure individuals, such as dietary changes chronic stress, and unhealthy coping mechanisms like smoking or reduced physical activity, can inform targeted interventions to effectively manage hypertension in vulnerable populations.

Our study supports prior research indicating a high prevalence of hypertension^26,27^ and food insecurity^28^ in Non-Hispanic Black participants. In contrast, Non-Hispanic White and Hispanic participants, despite having a high prevalence of food insecurity, did not exhibit a significant influence on hypertension risk. This emphasizes the complex interplay between ethnicity, food security, and health outcomes.^29^ The observed racial disparity in the influence of food insecurity on hypertension risk highlights the need for culturally sensitive interventions that address the broader SDoH can promote more equitable health outcomes.

Predictably, we found a higher prevalence of obesity, a known hypertension risk factor, in individuals from food-insecure households. Our finding that food insecure adults were less likely to be engaged in physical activity aligns with a study by To et al.^30^ This suggests a complex interplay wherein food insecurity may influence physical activity levels. The lack of access to healthy food or uncertainty in obtaining it may cause changes in physical activity behaviors, which in turn, may contribute to chronic diseases such as obesity and hypertension.^31^

It is important to acknowledge the study’s limitations. Firstly, potential selection bias may exist, given the response rate to the food security questionnaire and missing information in the dataset, affecting generalizability. Secondly, dietary assessment methods for sodium and potassium intake may be influenced by self-report, challenges in quantifying salt, and failure to account for non-food sources of sodium such as supplements, and nutrient coding errors. Thirdly, reliance on self-reported measures of food insecurity and dietary intake introduces potential recall bias or social desirability bias.^32^ Lastly, the cross-sectional design of NHANES limits causal inference, and the use of within-person mean instead of using generalized mixed methods for estimating nutrient intake simplifies the approach but may be more susceptible to errors.^33^

This study underscores the complex interplay between food insecurity, dietary habits, and hypertension. It highlights the potential need for interventions to improve food security and promote access to nutrient-rich foods to reduce hypertension risk. This emphasizes the need for policy changes addressing food access issues; ensuring available food resources include potassium-rich foods like fruits and vegetables. Efforts should focus on increasing access to affordable, healthy food options and expanding food assistance programs in low-income areas.^34^ Further research is warranted to comprehensively understand the complex mechanisms involved and develop strategies to enhance food security, improve dietary habits, and ultimately decrease hypertension risk among vulnerable populations.

In conclusion, the study underscores the critical role of SDoH, particularly food insecurity, in the prevalence of hypertension. Implementing policies and interventions to mitigate food insecurity and enhance dietary quality, especially in low-income communities, can significantly contribute to lessening the burden of hypertension.^35^

### Perspectives

In this nationally representative study, adults with food insecurity had reduced dietary potassium intake, with no significant association with average sodium intake based on food security status. Food insecurity, a modifiable risk factor for CVD, presents a pathway to decrease the burden of cardiovascular morbidity and mortality. The study reveals the impact of food insecurity on obesity and physical activity, both hypertension risk factors. Racial disparities in the impact of food insecurity on hypertension risk were suggested, warranting further research to explore this interaction. Our findings carry significant public health implications, emphasizing the importance of addressing food insecurity to reduce hypertension risk.

## NOVELTY AND RELEVANCE

**What Is New?**

This is the first study to evaluate the association of dietary patterns of sodium and potassium based on food security status on hypertension risk using a nationally representative sample.

**What Is Relevant?**

Food insecure participants had a lower dietary potassium intake compared to individuals who were food secure, likely related to lower intake of potassium-rich foods such as fruits, vegetables and grain, a known hypertension risk.

**Clinical/Pathophysiological Implications?**

Our findings highlight the critical impact of food insecurity on the prevalence of hypertension. We provide evidence to support interventions to reduce food insecurity and enhance dietary quality, which might contribute to decreasing the burden of hypertension.

## Data Availability

The NHANES data used is publically available

---

**Table S1.**
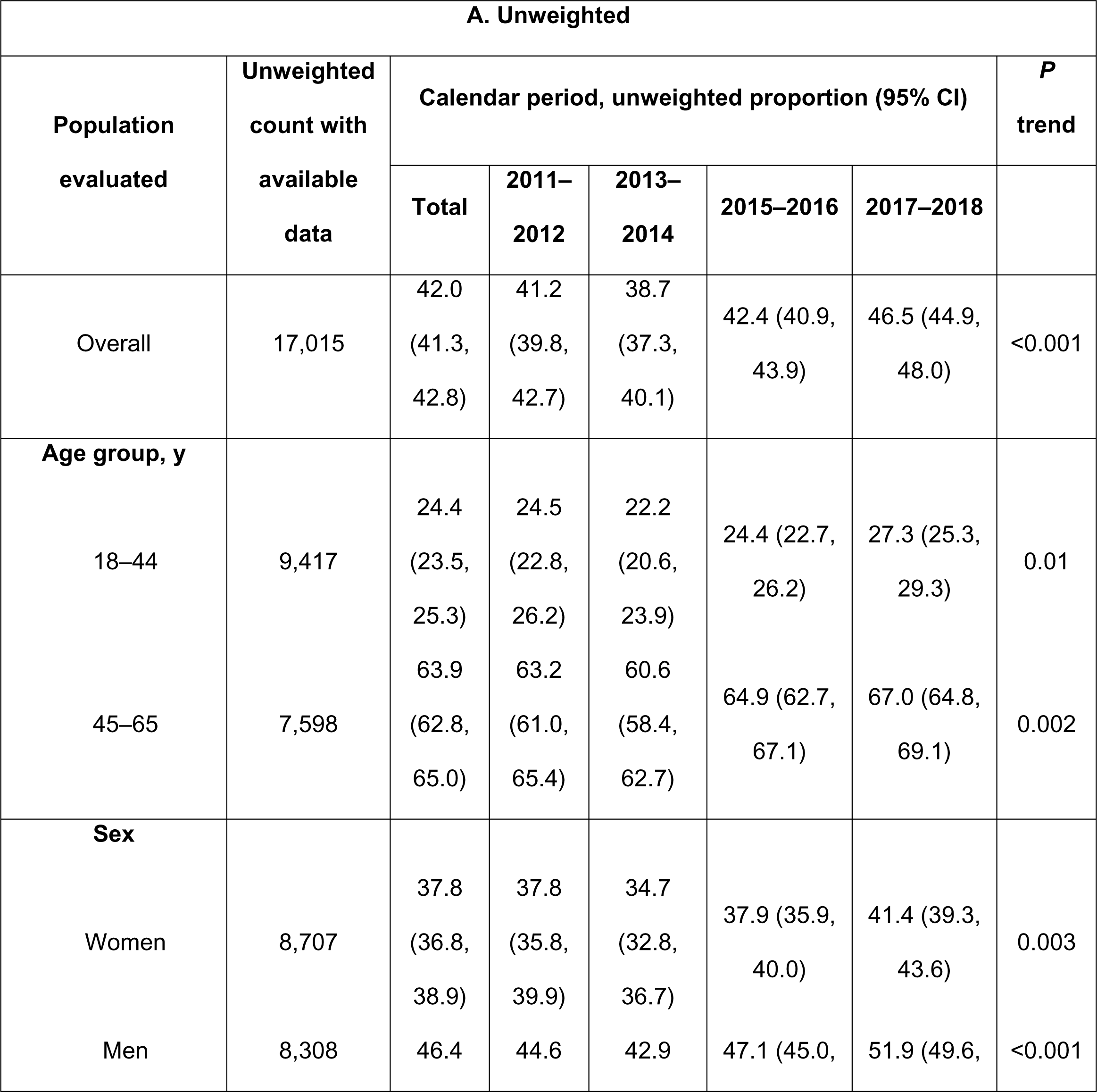
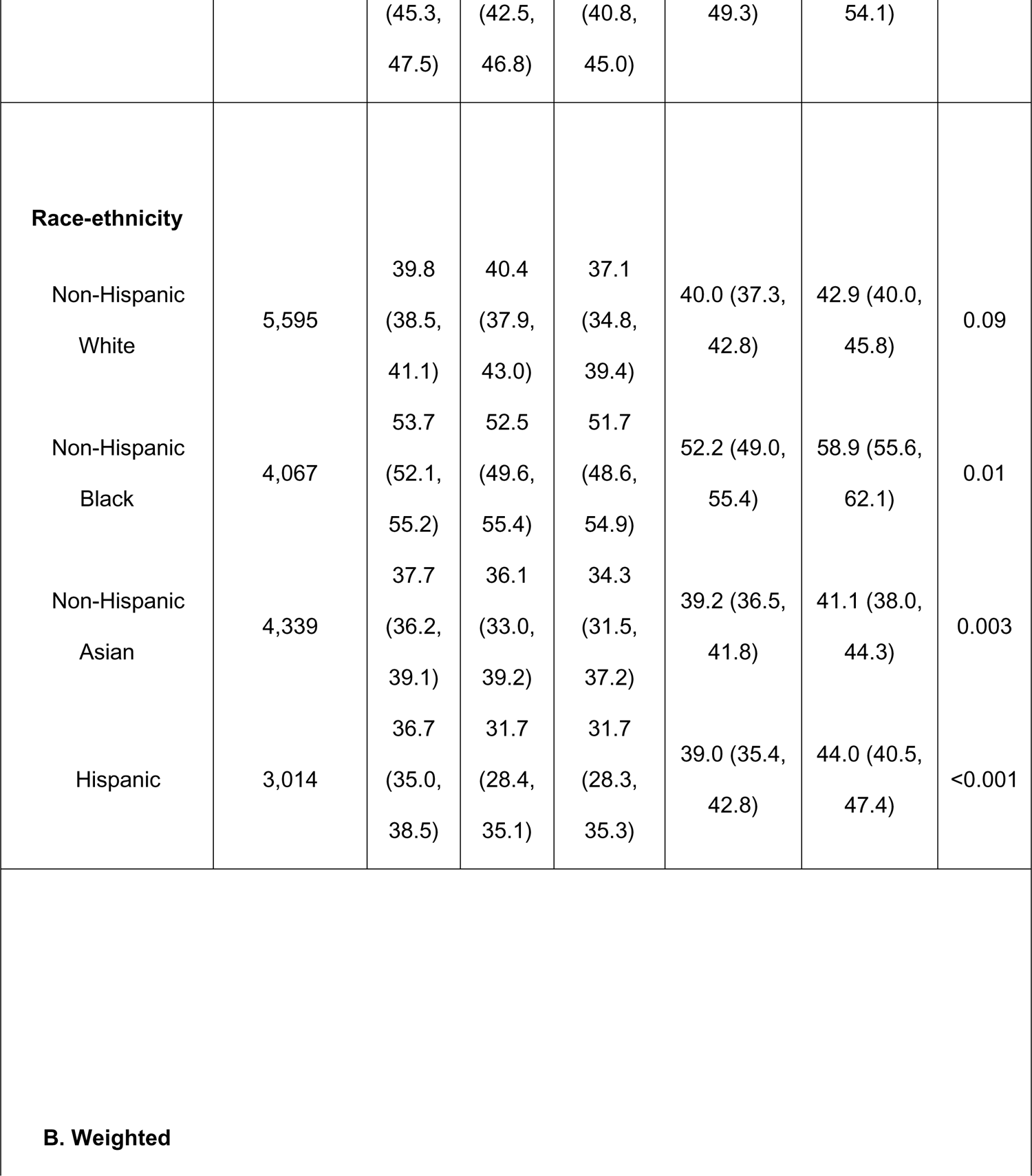
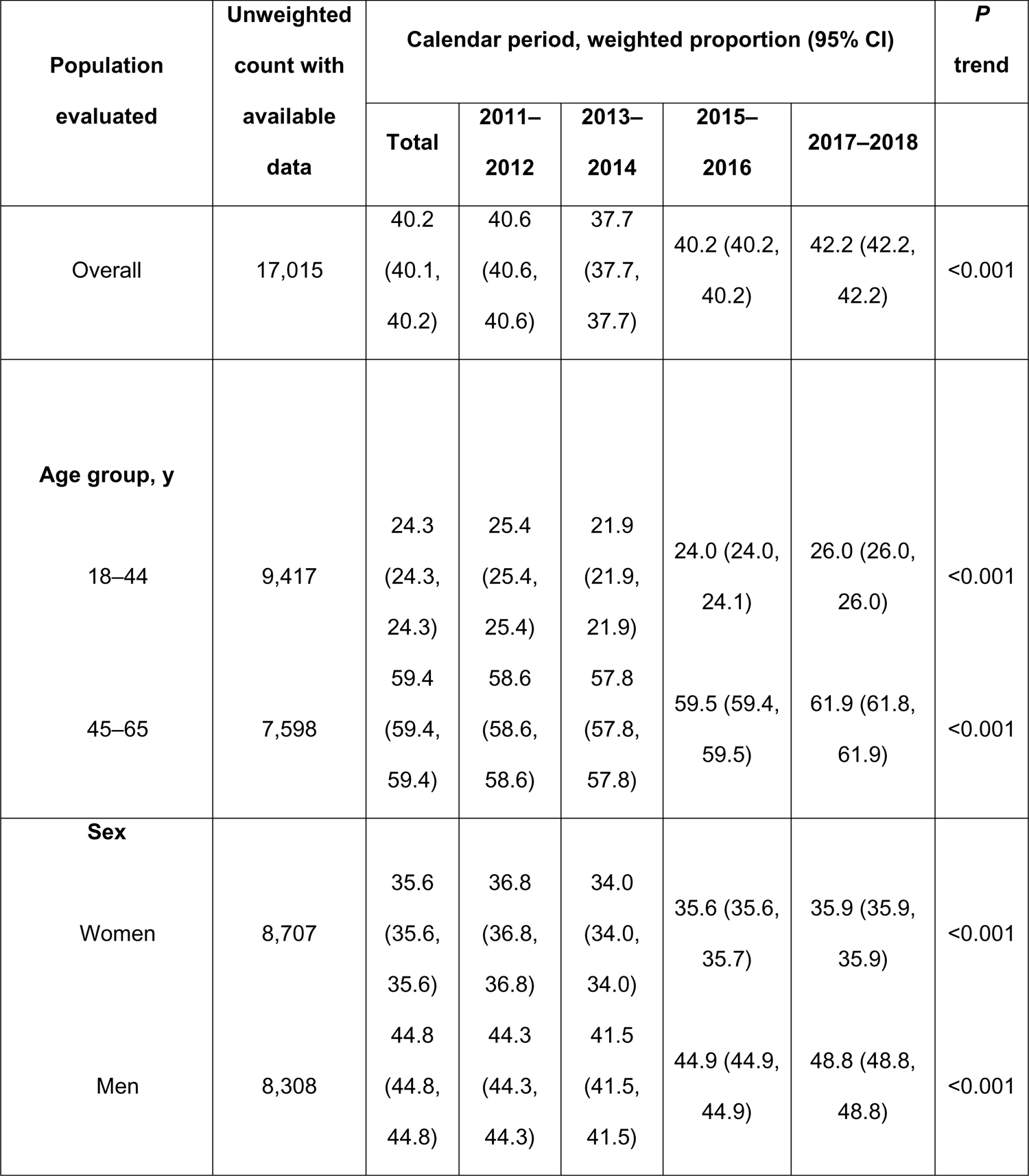
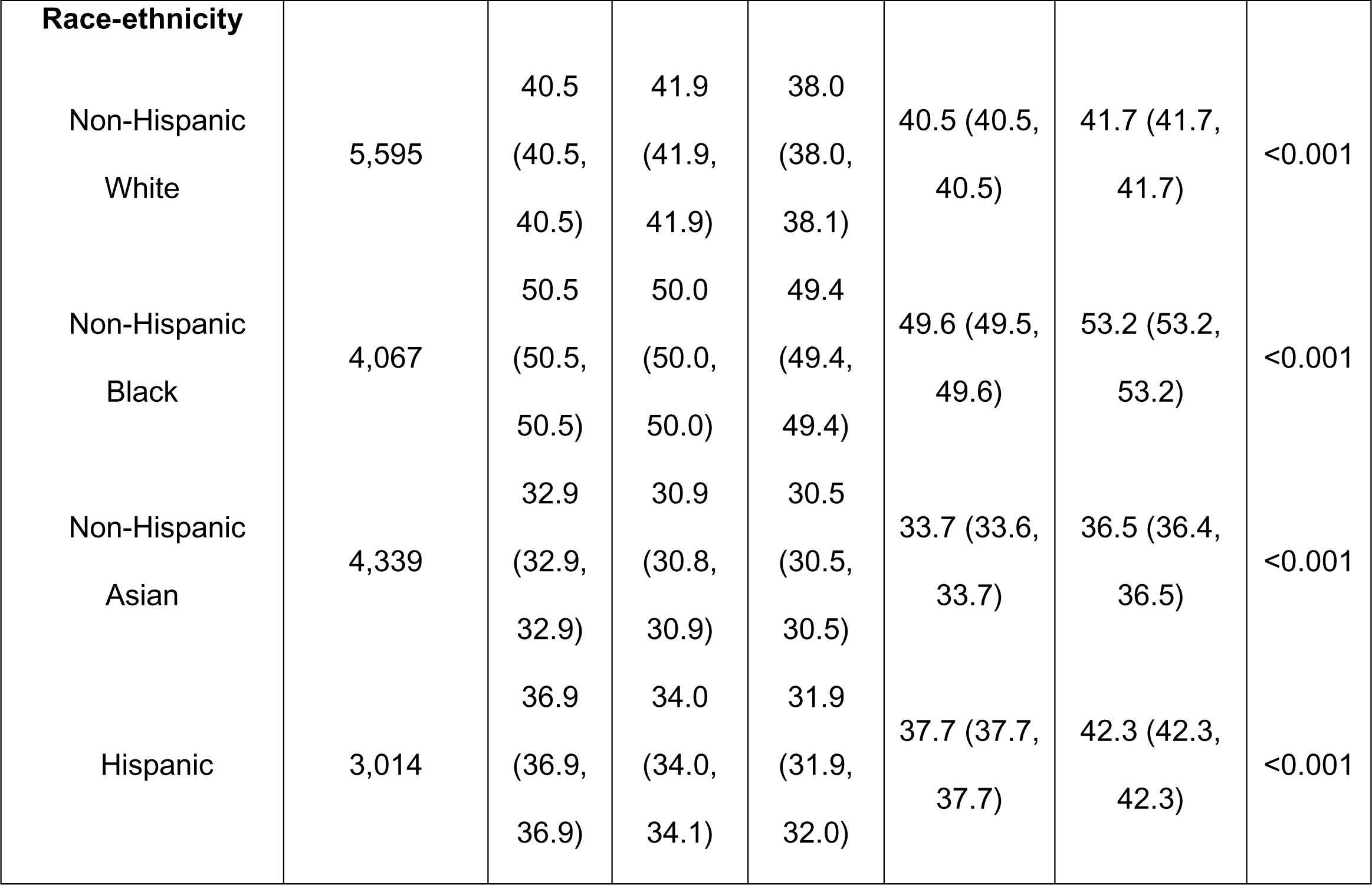
Age-Specific and Age-Adjusted Prevalence of Hypertension Among US Adults in 2011 to 2012, 2013 to 2016, and 2017 to 2018, overall and in Age, Sex, and Race-Ethnicity Subgroups

## References

1. Centers for Disease Control and Prevention. Estimated Hypertension Prevalence, Treatment, and Control Among U.S. Adults, https://millionhearts.hhs.gov/data-reports/hypertension-prevalence.html.

2. Schultz WM, Kelli HM, Lisko JC, et al. Socioeconomic Status and Cardiovascular Outcomes: Challenges and Interventions. Circulation 2018; 137: 2166–2178. 2018/05/16. DOI: 10.1161/CIRCULATIONAHA.117.029652.

3. Shahu A, Herrin J, Dhruva SS, et al. Disparities in Socioeconomic Context and Association With Blood Pressure Control and Cardiovascular Outcomes in ALLHAT. J Am Heart Assoc 2019; 8: e012277. 2019/08/01. DOI: 10.1161/JAHA.119.012277.

4. ERS. Food security in the US, https://www.ers.usda.gov/topics/food-nutrition-assistance/food-security-in-the-u-s/key-statistics-graphics/#foodsecure.

5. Nagata JM, Palar K, Gooding HC, et al. Food Insecurity and Chronic Disease in US Young Adults: Findings from the National Longitudinal Study of Adolescent to Adult Health. J Gen Intern Med 2019; 34: 2756–2762. 2019/10/03. DOI: 10.1007/s11606-019-05317-8.

6. Feeding America. Hunger in America: Executive Summary. 2014.

7. Liu Y, Eicher-Miller HA. Food Insecurity and Cardiovascular Disease Risk. Curr Atheroscler Rep. 2021 Mar 27;23(6):24. doi: 10.1007/s11883-021-00923-6. PMID: 33772668; PMCID: PMC8000689.

8. Sacks FM, Svetkey LP, Vollmer WM, Appel LJ, et al; DASH-Sodium Collaborative Research Group. Effects on blood pressure of reduced dietary sodium and the Dietary Approaches to Stop Hypertension (DASH) diet. DASH-Sodium Collaborative Research Group. N Engl J Med. 2001 Jan 4;344(1):3–10. doi: 10.1056/NEJM200101043440101. PMID: 11136953.

9. Palakshappa D, Ip EH, Berkowitz SA, Bertoni AG, et al. Pathways by Which Food Insecurity Is Associated With Atherosclerotic Cardiovascular Disease Risk. J Am Heart Assoc. 2021 Nov 16;10(22):e021901. doi: 10.1161/JAHA.121.021901. Epub 2021 Nov 6. PMID: 34743567; PMCID: PMC8751929.

10. Seligman HK, Laraia BA, Kushel MB. Food insecurity is associated with chronic disease among low-income NHANES participants. J Nutr. 2010 Feb;140(2):304–10. doi: 10.3945/jn.109.112573. Epub 2009 Dec 23. Erratum in: J Nutr. 2011 Mar;141(3):542. PMID: 20032485; PMCID: PMC2806885.

11. Rasmusson G, Lydecker JA, Coffino JA, et al. Household food insecurity is associated with binge-eating disorder and obesity. Int J Eat Disord 2018 2018/12/20. DOI: 10.1002/eat.22990.

12. Leung CW, Epel ES, Ritchie LD, et al. Food insecurity is inversely associated with diet quality of lower-income adults. J Acad Nutr Diet 2014; 114: 1943–1953.e1942. DOI: 10.1016/j.jand.2014.06.353.

13. Brown AGM, Esposito LE, Fisher RA, Nicastro HL, et al. Food insecurity and obesity: research gaps, opportunities, and challenges. Transl Behav Med. 2019 Oct 1;9(5):980–987. doi: 10.1093/tbm/ibz117. PMID: 31570918; PMCID: PMC6937550.

14. Li M, Yan S, Li X, Jiang S, Ma X, et al. Association between blood pressure and dietary intakes of sodium and potassium among US adults using quantile regression analysis NHANES 2007-2014. J Hum Hypertens. 2020 May;34(5):346–354. doi: 10.1038/s41371-019-0224-9. Epub 2019 Aug 16. PMID: 31420581.

15. Gundersen C, Engelhard E and Hake M. The Determinants of Food Insecurity among Food Bank Clients in the United States. Journal of Consumer Affairs 2017; 51: 501–518. DOI: 10.1111/joca.12157.

16. National Center for Health Statistics. About the National Health and Nutrition Examination Survey, (January 25, 2024).

17. Whelton PK, Carey RM, Aronow WS, Casey DE Jr, et al. 2017 ACC/AHA/AAPA/ABC/ACPM/AGS/APhA/ASH/ASPC/NMA/PCNA Guideline for the Prevention, Detection, Evaluation, and Management of High Blood Pressure in Adults: A Report of the American College of Cardiology/American Heart Association Task Force on Clinical Practice Guidelines. Hypertension. 2018 Jun;71(6):e13–e115. doi: 10.1161/HYP.0000000000000065. Epub 2017 Nov 13. Erratum in: Hypertension. 2018 Jun;71(6):e140-e144. PMID: 29133356.

18. US Department of Agriculture. Survey Tools, https://www.ers.usda.gov/topics/food-nutrition-assistance/food-security-in-the-u-s/survey-tools/.

19. National Center for Health Statistics. National Health and Nutrition Examination Survey 2015–2016 Data Documentation, Codebook, and Frequencies. Available online: https://wwwn.cdc.gov/Nchs/Nhanes/2015-2016/FSQ_I.htm (accessed on 6 November 2023).

20. Bauman A, Phongsavan P, Schoeppe S, Owen N. Physical activity measurement--a primer for health promotion. Promot Educ. 2006;13(2):92–103. doi: 10.1177/10253823060130020103. PMID: 17017286.

21. Diaz T, Strong KL, Cao B, Guthold R, et al. A call for standardised age-disaggregated health data. Lancet Healthy Longev. 2021 Jul;2(7):e436–e443. doi: 10.1016/S2666-7568(21)00115-X. Erratum in: Lancet Healthy Longev. 2021 Aug;2(8):e458. PMID: 34240065; PMCID: PMC8245325.

22. Micha R, Peñalvo JL, Cudhea F, Imamura F, et al. Association Between Dietary Factors and Mortality From Heart Disease, Stroke, and Type 2 Diabetes in the United States. JAMA. 2017 Mar 7;317(9):912–924. doi: 10.1001/jama.2017.0947. PMID: 28267855; PMCID: PMC5852674.

23. Elmadfa I, Meyer AL. Importance of food composition data to nutrition and public health. Eur J Clin Nutr. 2010 Nov;64 Suppl 3:S4–7. doi: 10.1038/ejcn.2010.202. PMID: 21045848.

24. Beltrán S, Pharel M, Montgomery CT, López-Hinojosa IJ, et al. Food insecurity and hypertension: A systematic review and meta-analysis. PLoS One. 2020 Nov 17;15(11):e0241628. doi: 10.1371/journal.pone.0241628. PMID: 33201873; PMCID: PMC7671545.

25. Ing CT, Clemens B, Ahn HJ, Kaholokula JK, et al. Food Insecurity and Blood Pressure in a Multiethnic Population. Int J Environ Res Public Health. 2023 Jun 28;20(13):6242. doi: 10.3390/ijerph20136242. PMID: 37444090; PMCID: PMC10341426.

26. Hertz RP, Unger AN, Cornell JA, Saunders E. Racial disparities in hypertension prevalence, awareness, and management. Arch Intern Med. 2005 Oct 10;165(18):2098–104. doi: 10.1001/archinte.165.18.2098. PMID: 16216999.

27. Aggarwal R, Chiu N, Wadhera RK, Moran AE, et al. Racial/Ethnic Disparities in Hypertension Prevalence, Awareness, Treatment, and Control in the United States, 2013 to 2018. Hypertension. 2021 Dec;78(6):1719–1726. doi:10.1161/HYPERTENSIONAHA.121.17570. Epub 2021 Aug 9. PMID: 34365809.

28. Dhunna S, Tarasuk V. Black-white racial disparities in household food insecurity from 2005 to 2014, Canada. Can J Public Health. 2021 Oct;112(5):888–902. doi: 10.17269/s41997-021-00539-y. Epub 2021 Jun 15. PMID: 34129216; PMCID: PMC8204605.

29. Fernandez J, García-Pérez M and Orozco-Aleman S. Unraveling the Hispanic Health Paradox. Journal of Economic Perspectives 2023; 37: 145–167. DOI: 10.1257/jep.37.1.145.

30. To QG, Frongillo EA, Gallegos D, et al. Household food insecurity is associated with less physical activity among children and adults in the U.S. population. J Nutr 2014; 144: 1797–1802. 2014/10/22. DOI: 10.3945/jn.114.198184.

31. Bateson M, Pepper GV. Food insecurity as a cause of adiposity: evolutionary and mechanistic hypotheses. Philos Trans R Soc Lond B Biol Sci. 2023 Oct 23;378(1888):20220228. doi: 10.1098/rstb.2022.0228. Epub 2023 Sep 4. PMID: 37661744; PMCID: PMC10475876.

32. Hager ER, Quigg AM, Black MM, et al. Development and validity of a 2-item screen to identify families at risk for food insecurity. Pediatrics 2010; 126: e26–32. 2010/07/03. DOI: 10.1542/peds.2009-3146.

33. Herrick KAR, L.M.; Parsons, R.; Dodd,K.W. Estimating usual dietary intake from National Health and Nutrition Examination Survey data using the National Cancer Institute method. In: Statistics NCfH, (ed.). 2018.

34. Gittelsohn J, Kasprzak CM, Hill AB, Sundermeir SM, et al. Increasing Healthy Food Access for Low-Income Communities: Protocol of the Healthy Community Stores Case Study Project. Int J Environ Res Public Health. 2022 Jan 8;19(2):690. doi: 10.3390/ijerph19020690. PMID: 35055512; PMCID: PMC8775718.

35. Ziso D, Chun OK, Puglisi MJ. Increasing Access to Healthy Foods through Improving Food Environment: A Review of Mixed Methods Intervention Studies with Residents of Low-Income Communities. Nutrients. 2022 May 29;14(11):2278. doi: 10.3390/nu14112278. PMID: 35684077; PMCID: PMC9182982.

